# An Empirical Inference of the Severity of Resurgence of COVID-19 in Europe

**DOI:** 10.1101/2020.10.21.20213868

**Authors:** Hemanta K. Baruah

**Affiliations:** Department of Mathematics, The Assam Royal Global University, Guwahati, Assam, India

**Keywords:** Pandemic, infectious disease, epidemiological modeling

## Abstract

In Europe the Corona Virus spread had started to retard months ago, but after some time it has started to accelerate again. In this article, we are going to analyze the current COVID-19 spread patterns in Italy, the UK, Germany, Russia, Spain and France. We have found that the current spread has perhaps been underestimated as just the second wave. As per our analysis, as on 7 October the resurgence is much more vigorous than the first wave of spread of the disease. It is going to be most serious in Russia, followed by Italy, Germany and the UK, while in Spain and France the patterns are yet to take inferable shapes.

## Introduction

In the European countries the second wave of COVID-19 has been observed to be much more serious than the first one. The data clearly reflect resurgence of the disease, but how serious the matter is can be judged only after a proper data analysis. In this article, we are going to show how to infer empirically the severity of the spread. We shall show that the COVID-19 situation in Europe is indeed very serious.

A look at the daily increase in the cumulative totals of the COVID-19 cases in the European countries mentioned above and the World gives us an apparently clear picture how much Europe is contributing to the daily increase in the total number of cases in the World. For example, on October 7 the increase in the World total were 348967 out of which the UK contributed 14162 (4.06%) cases, Italy contributed 3678 (1.11%) cases, Germany contributed 3994 (1.14%) cases, France contributed 18746 (5.37%) cases, Russia contributed 11115 (3.18%) cases and Spain contributed 10574 (3.03%) cases. The total increase on that day in these six European countries were 62269 (17.84%) to the total increase in the World. On that same day, India which is currently contributing the maximum to the daily increase in the World contributed 78809 (22.58%) cases while the USA on that day contributed 49398 (14.15%) cases. Therefore it can be seen that on October 7, these six European countries together had contributed more to the World total than the USA although the contribution was less than that from India. However, we have to note that in India and the USA, the spread is retarding while in these six European countries that kind of retardations took place months ago, and thereafter it has started to move towards resurgence – the so called second wave. Indeed the figures for one single day may actually be misleading. We shall put forward a numerical analysis of the spread data for a period of 15 days from 23 September to 7 October.

We would like to mention at this point that the current growth curves of the disease in these six European countries are obviously not like the curve assumed in the compartmental epidemiological models [1, 2. 3]. At the start of an epidemic, the classical compartmental models do not presume a change from a retarding state to an accelerating state.

## Methodology

It is apparent from the graphs published by Worldometers.info [4] that the spread patterns are of the exponential type in the six European countries mentioned above. We shall consider the total number of cases in these six countries from 22 September to 7 October. From that we shall form a time series from 23 September to 7 October, of the values of Δ*z*(*t*) for t = 1, 2, …, 15, where Δ*z*(*t*) are the first order differences of

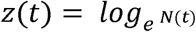

where *N*(*t*) is the cumulative total number of cases at time *t*. We shall first see whether the spread is approximately following the function

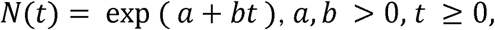

by the observations. We shall thereafter demonstrate diagrammatically that the time series of Δ*z*(*t*) would explain what exactly the picture is as far as resurgence of the disease is concerned.

Indeed for *N*(*t*) strictly exponential following the pattern shown above Δ*z*(*t*) would be constant. But in the case of an epidemic, the function *N*(*t*) cannot be strictly exponential. It can be only approximately exponential. It was earlier observed that for Δ*t* = 1, Δ*z*(*t*) follows the following rules. Once the cumulative total number of cases of the epidemic had entered into the nearly exponential phase, Δ*z*(*t*) starts to follow a reducing trend. It was earlier observed that a change from a nearly exponential pattern to a nearly logarithmic pattern took place in Italy [5] months ago. In the other five European countries too such retardation must have followed the nearly logarithmic pattern. It was observed [6, 7, 8, 9, 10] that when the pandemic continues to grow nearly exponentially, Δ*z*(*t*) would continue to decrease linearly in time. If it continues to be very nearly constant for a sufficiently long duration after continuing to decrease linearly earlier, that should be taken as a signal that the pattern might start to change to a nearly logarithmic one soon after. It was shown in [11] how Δ*z*(*t*) can lead to conclude about the current situations in India, in the USA and in the World. Data from 17 September to 1 October were used. It was observed that the values of Δ*z*(*t*) were decreasing steadily in India, and were very nearly constants in the USA and the World. Based on the current procedure using data from 17 September to 1 October, forecasts regarding the cumulative total number of cases from 2 October to 16 October were made for India, the USA and the World.

It can be seen how close the forecasts [11] and the observed values are for the USA and the World. For India, it was mentioned that as in the forecasts the average of Δ*z*(*t*) was used, the forecasts would be overestimations. In effect, it was demonstrated that the method of inferring from the values of Δ*z*(*t*) is really useful. We now proceed to analyze the recent data on the cumulative total number of COVID-19 cases in the six European countries mentioned above.

## Analysis

We shall now put forward our empirical analysis of the COVI-19 spread patterns in the six European countries concerned. In Table-1 we have shown the values of Δ*z*(*t*) from 7 October downwards to 23 September for Italy. The values of Δ*z*(*t*) from 23 September to 7 October have been shown diagrammatically in Fig. 1.

**Table-1:** Values of Δ*z*(*t*), Italy, 23 September to 7 October.

| Date | $N(t)$ | $z(t)$ | $\Delta z(t)$ |
| --- | --- | --- | --- |
| 7-Oct | 333940 | 12.71872 | 0.011075 |
| 6-Oct | 330262 | 12.70764 | 0.008136 |
| 5-Oct | 327586 | 12.69951 | 0.006914 |
| 4-Oct | 325329 | 12.69259 | 0.007956 |
| 3-Oct | 322751 | 12.68464 | 0.008851 |
| 2-Oct | 319907 | 12.67579 | 0.007839 |
| 1-Oct | 317409 | 12.66795 | 0.00806 |
| 30-Sep | 314861 | 12.65989 | 0.005896 |
| 29-Sep | 313010 | 12.65399 | 0.005276 |
| 28-Sep | 311363 | 12.64871 | 0.004807 |
| 27-Sep | 309870 | 12.64391 | 0.005715 |
| 26-Sep | 308104 | 12.63819 | 0.006085 |
| 25-Sep | 306235 | 12.63211 | 0.006263 |
| 24-Sep | 304323 | 12.62584 | 0.005886 |
| 23-Sep | 302537 | 12.61996 | 0.005436 |
| 22-Sep | 300897 | 12.61452 | ---- |

**Fig. 1:**
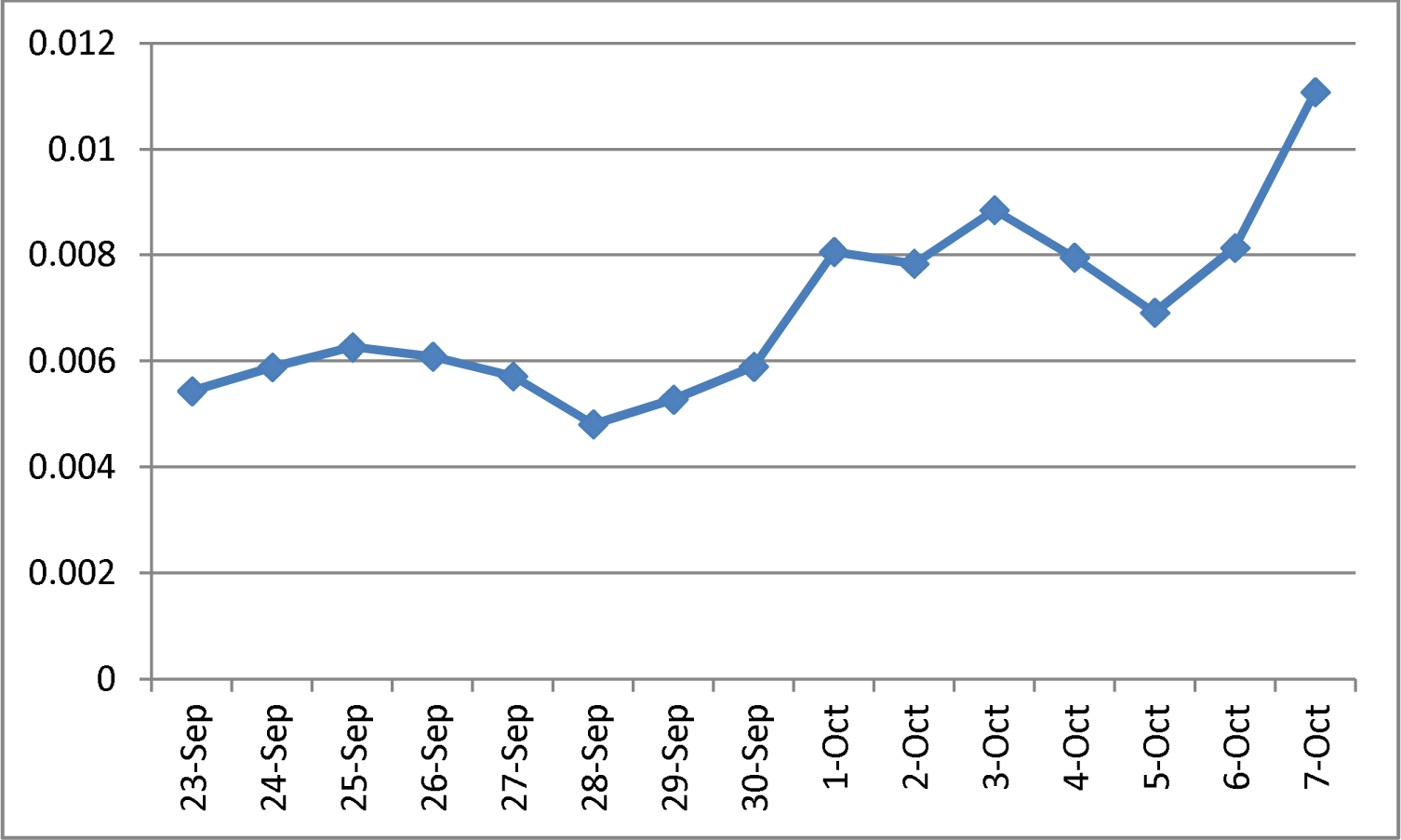
Values of Δ*z*(*t*), Italy, 23 September to 7 October.

From the diagrammatic representation, it is clear that what has been happening in Italy is unusual. The values of Δ*z*(*t*) are following an increasing trend around the average 0.006946. Indeed this should not have been the case. As we have mentioned earlier, in the nearly exponential phase, the values of Δ*z*(*t*) decreases linearly, and by the time the retardation starts the values of Δ*z*(*t*) start to become nearly constant. But currently in Italy the values of Δ*z*(*t*) have been found to be increasing. This gives us a picture that the COVID-19 situation in Italy is showing an unusual increasing trend, and such a thing is not assumed in any classical epidemiological mathematical model.

In Table-2, the values of Δ*z*(*t*) have been shown for the UK from 7 October downwards to 23 September. The values of Δ*z*(*t*) from 23 September to 7 October have been shown diagrammatically in Fig. 2. As can be seen, just like in Italy, in the UK too the values of Δ*z*(*t*) are showing an unusual increasing trend around the average 0.019849.

**Table-2:** Values of Δ*z*(*t*), the UK, 23 September to 7 October.

| Date | $N(t)$ | $z(t)$ | $\Delta z(t)$ |
| --- | --- | --- | --- |
| 7-Oct | 544275 | 13.20721 | 0.026364 |
| 6-Oct | 530113 | 13.18085 | 0.027815 |
| 5-Oct | 515571 | 13.15303 | 0.024731 |
| 4-Oct | 502977 | 13.1283 | 0.015995 |
| 3-Oct | 494996 | 13.1123 | 0.014386 |
| 2-Oct | 487926 | 13.09792 | 0.024385 |
| 1-Oct | 476172 | 13.07353 | 0.023473 |
| 30-Sep | 465125 | 13.05006 | 0.022079 |
| 29-Sep | 454968 | 13.02798 | 0.02024 |
| 28-Sep | 445852 | 13.00774 | 0.010511 |
| 27-Sep | 441190 | 12.99723 | 0.017637 |
| 26-Sep | 433477 | 12.97959 | 0.017639 |
| 25-Sep | 425898 | 12.96196 | 0.01781 |
| 24-Sep | 418380 | 12.94415 | 0.017741 |
| 23-Sep | 411023 | 12.9264 | 0.016927 |
| 22-Sep | 404124 | 12.90948 | ---- |

**Fig. 2:**
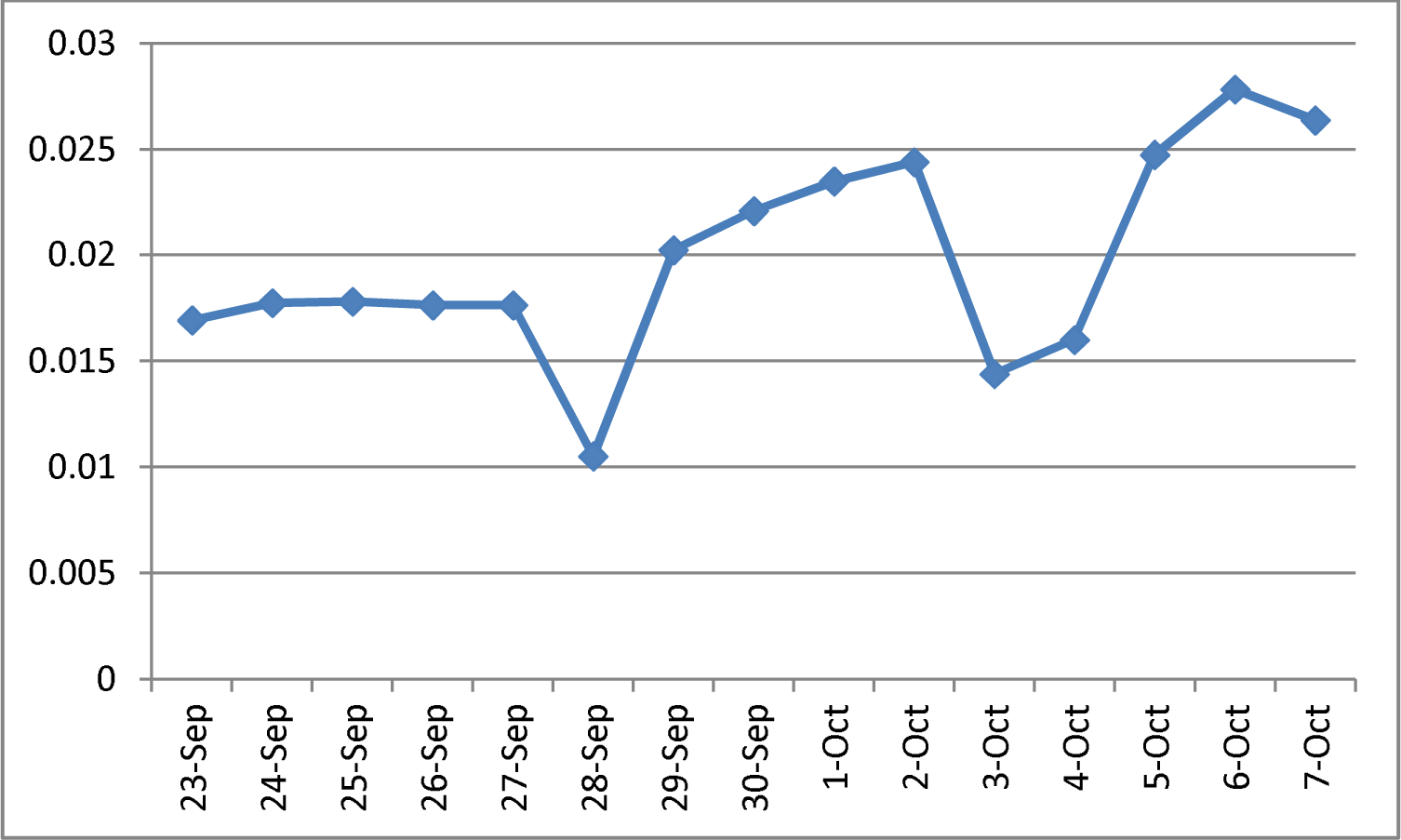
Values of Δ*z*(*t*), the UK, 23 September to 7 October.

In Table-3, the values of Δ*z*(*t*) have been shown for Germany from 7 October downwards to 23 September. The values of Δ*z*(*t*) from 23 September to 7 October have been shown diagrammatically in Fig. 3. As can be seen, just like in Italy and in the UK, in Germany too the values of Δ*z*(*t*) are showing an unusual increasing trend around the average 0.0077.

**Table-3:**
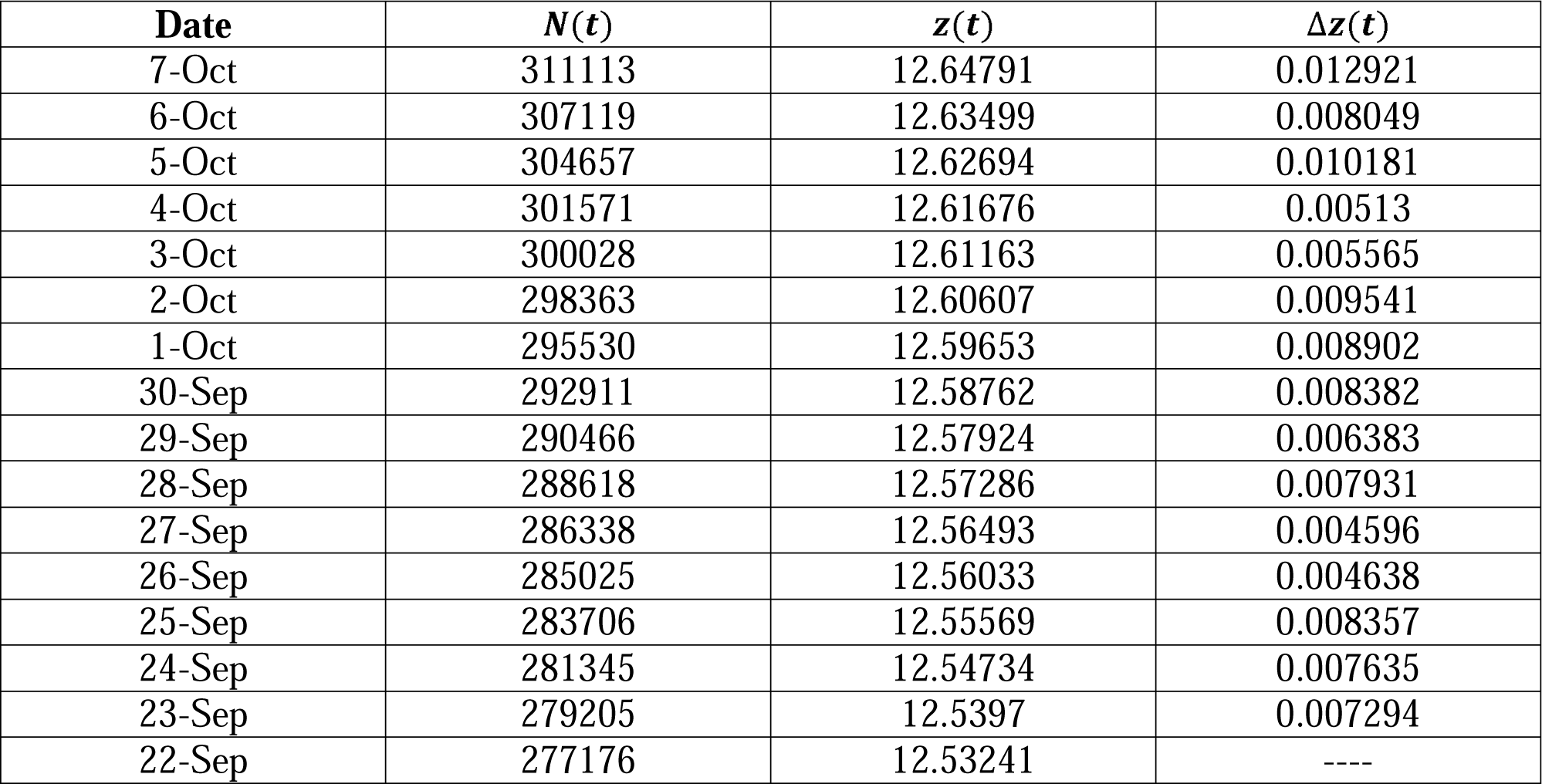
Values of Δ*z*(*t*), Germany, 23 September to 7 October.

**Fig. 3:**
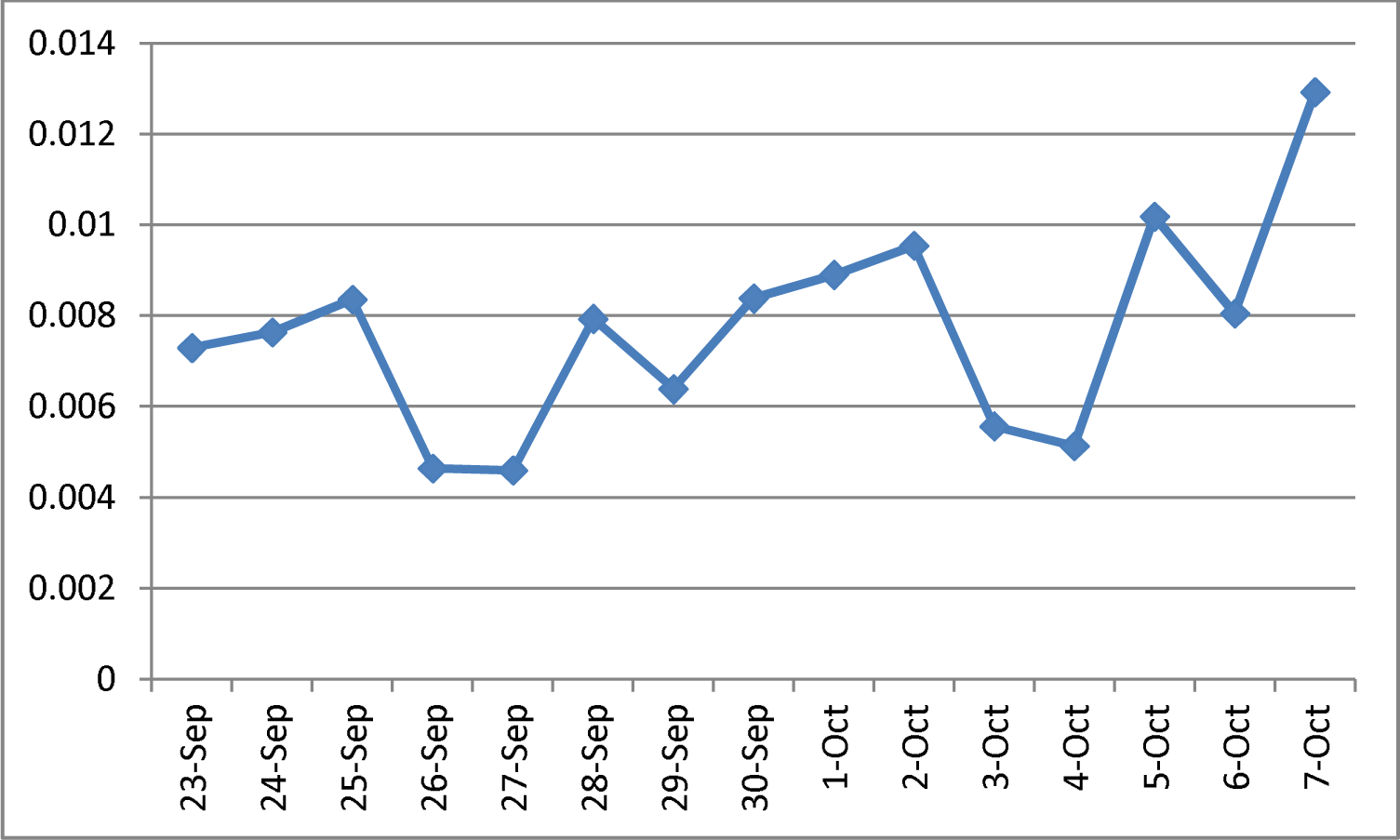
Values of Δ*z*(*t*), Germany, 23 September to 7 October.

In Russia, Table-4, the values of Δ*z*(*t*) have shown a straight increasing trend which is very unusual in the growth of an epidemic. How long it would continue to have such an increasing trend is not possible to judge. A diagrammatic representation has been shown in Fig. 4.

**Table-4:** Values of Δ*z*(*t*), Russia, 23 September to 7 October.

| Date | $N(t)$ | $z(t)$ | $\Delta z(t)$ |
| --- | --- | --- | --- |
| 7-Oct | 1248149 | 14.03717 | 0.008565 |
| 6-Oct | 1237504 | 14.02861 | 0.00943 |
| 5-Oct | 1225889 | 14.01918 | 0.008921 |
| 4-Oct | 1215001 | 14.01026 | 0.008679 |
| 3-Oct | 1204502 | 14.00158 | 0.008219 |
| 2-Oct | 1194643 | 13.99336 | 0.00791 |
| 1-Oct | 1185231 | 13.98545 | 0.007576 |
| 30-Sep | 1176286 | 13.97787 | 0.007236 |
| 29-Sep | 1167805 | 13.97064 | 0.007074 |
| 28-Sep | 1159573 | 13.96356 | 0.00704 |
| 27-Sep | 1151438 | 13.95652 | 0.006856 |
| 26-Sep | 1143571 | 13.94967 | 0.0066 |
| 25-Sep | 1136048 | 13.94307 | 0.006369 |
| 24-Sep | 1128836 | 13.9367 | 0.005859 |
| 23-Sep | 1122241 | 13.93084 | 0.005747 |
| 22-Sep | 1115810 | 13.92509 | ---- |

**Fig. 4:**
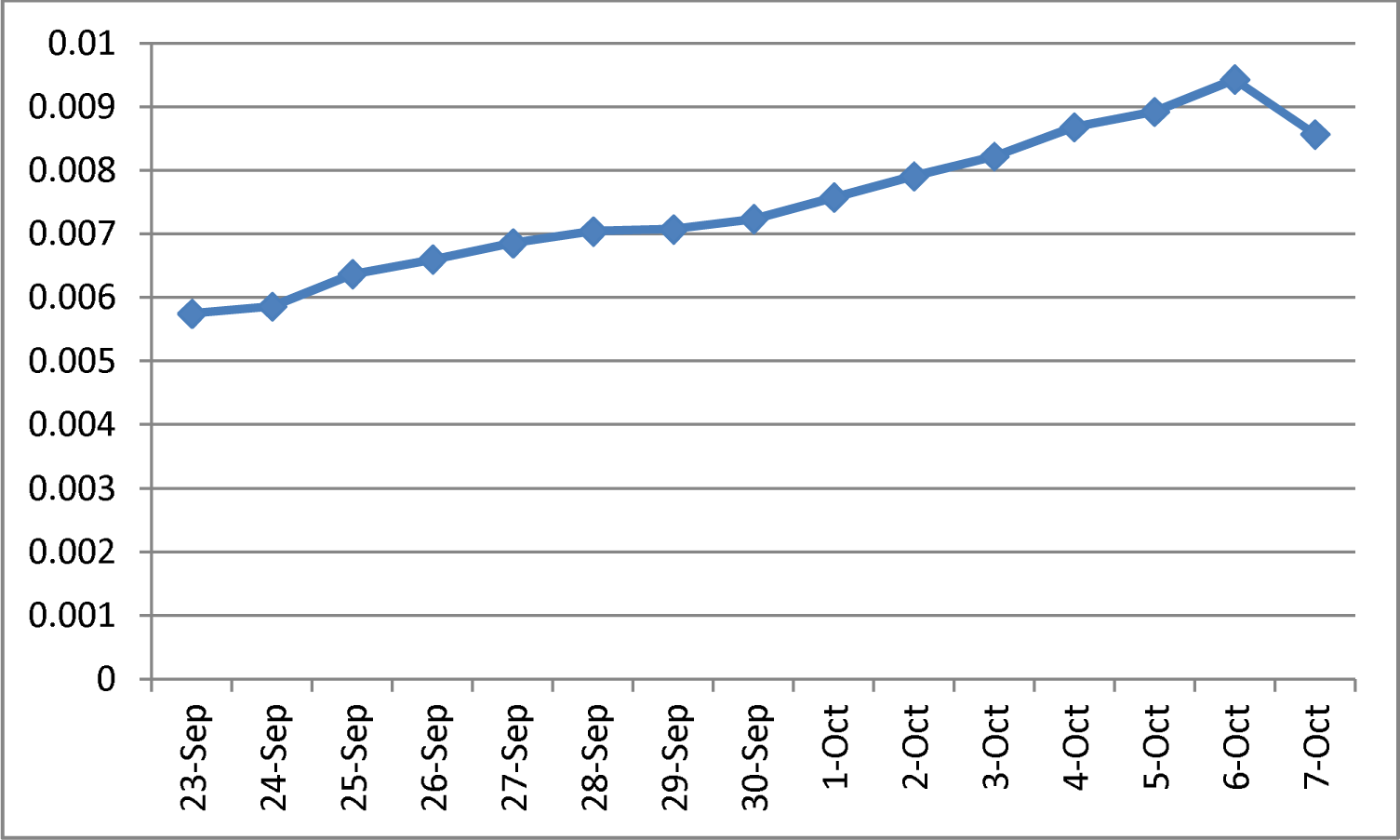
Values of Δ*z*(*t*), Russia, 23 September to 7 October.

In Spain, Table-5, the situation is different from that in Italy, the UK, Germany and Russia. In Spain, no increasing trend could be seen, but the values are not showing any constancy and they are not showing any decreasing trend as well. The diagrammatic representation has been shown in Fig. 5.

**Table-5:** Values of Δ*z*(*t*), Spain, 23 September to 7 October.

| Date | $N(t)$ | $z(t)$ | $\Delta z(t)$ |
| --- | --- | --- | --- |
| 7-Oct | 895503 | 13.70514 | 0.011878 |
| 6-Oct | 884929 | 13.69326 | 0.012919 |
| 5-Oct | 873570 | 13.68034 | 0.011261 |
| 4-Oct | 863788 | 13.66908 | 0.007139 |
| 3-Oct | 857643 | 13.66194 | 0.007938 |
| 2-Oct | 850862 | 13.65401 | 0.012188 |
| 1-Oct | 840555 | 13.64182 | 0.011212 |
| 30-Sep | 831183 | 13.63061 | 0.011256 |
| 29-Sep | 821880 | 13.61935 | 0.012051 |
| 28-Sep | 812035 | 13.6073 | 0.010363 |
| 27-Sep | 803663 | 13.59694 | 0.00738 |
| 26-Sep | 797754 | 13.58956 | 0.009262 |
| 25-Sep | 790399 | 13.58029 | 0.014105 |
| 24-Sep | 779329 | 13.56619 | 0.013924 |
| 23-Sep | 768553 | 13.55226 | 0.015113 |
| 22-Sep | 757025 | 13.53715 | ---- |

**Fig. 5:**
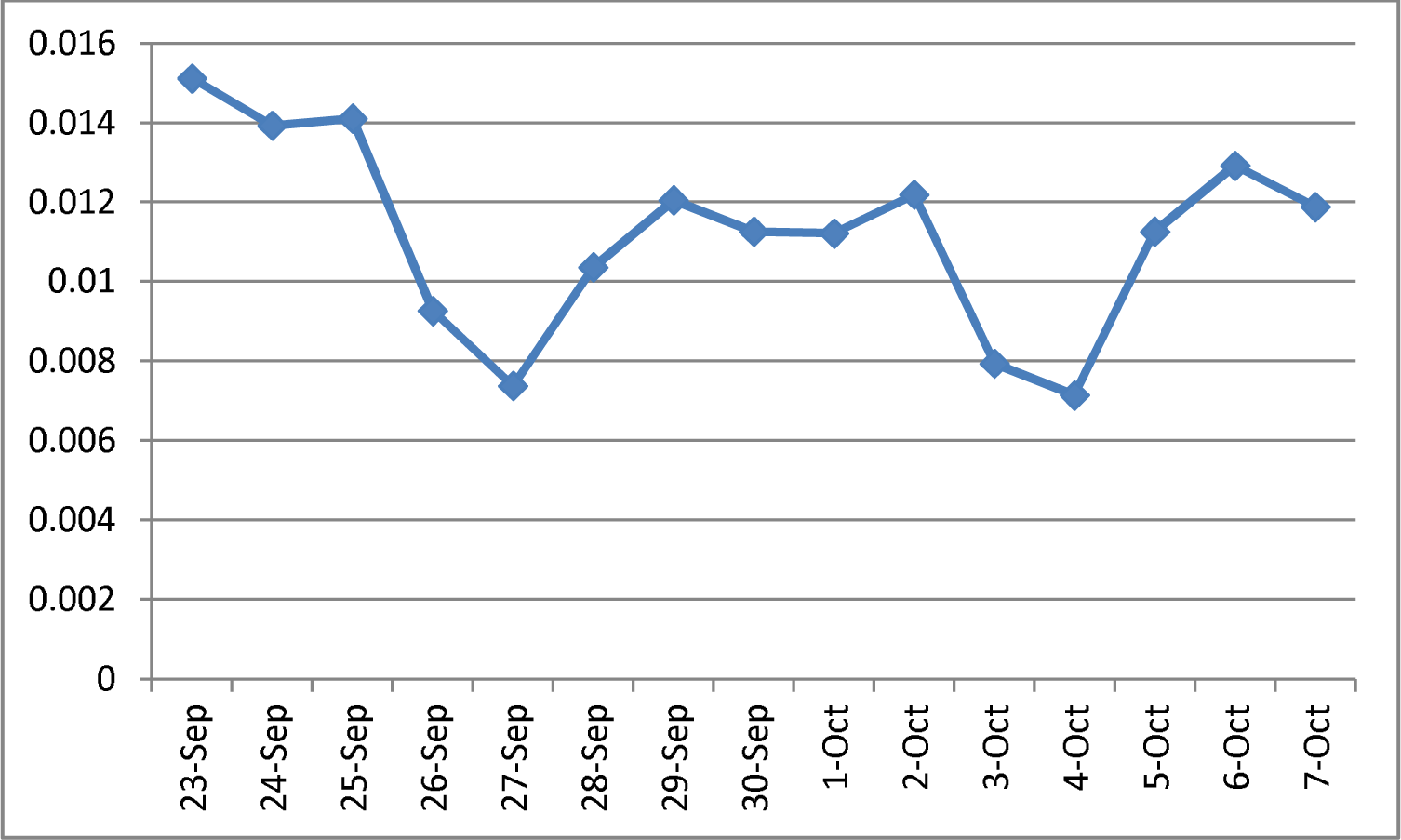
Values of Δ*z*(*t*), Spain, 23 September to 7 October.

In France too, Table-6, just like in Spain the values of Δ*z*(*t*) have been found to be nowhere near any constant value. The values are not increasing, but they are not showing any decreasing trend also. The diagrammatic representation has been shown in Fig. 6.

**Table-6:** Values of Δ*z*(*t*), France, 23 September to 7 October.

| Date | $N(t)$ | $z(t)$ | $\Delta z(t)$ |
| --- | --- | --- | --- |
| 7-Oct | 653509 | 13.39011 | 0.029105 |
| 6-Oct | 634763 | 13.36101 | 0.016662 |
| 5-Oct | 624274 | 13.34434 | 0.008177 |
| 4-Oct | 619190 | 13.33617 | 0.020501 |
| 3-Oct | 606625 | 13.31567 | 0.028377 |
| 2-Oct | 589653 | 13.28729 | 0.020817 |
| 1-Oct | 577505 | 13.26647 | 0.024488 |
| 30-Sep | 563535 | 13.24198 | 0.023057 |
| 29-Sep | 550690 | 13.21893 | 0.014728 |
| 28-Sep | 542639 | 13.2042 | 0.007529 |
| 27-Sep | 538569 | 13.19667 | 0.020869 |
| 26-Sep | 527446 | 13.1758 | 0.027704 |
| 25-Sep | 513034 | 13.1481 | 0.031275 |
| 24-Sep | 497237 | 13.11682 | 0.032906 |
| 23-Sep | 481141 | 13.08392 | 0.027545 |
| 22-Sep | 468069 | 13.05637 | ---- |

**Fig. 6:**
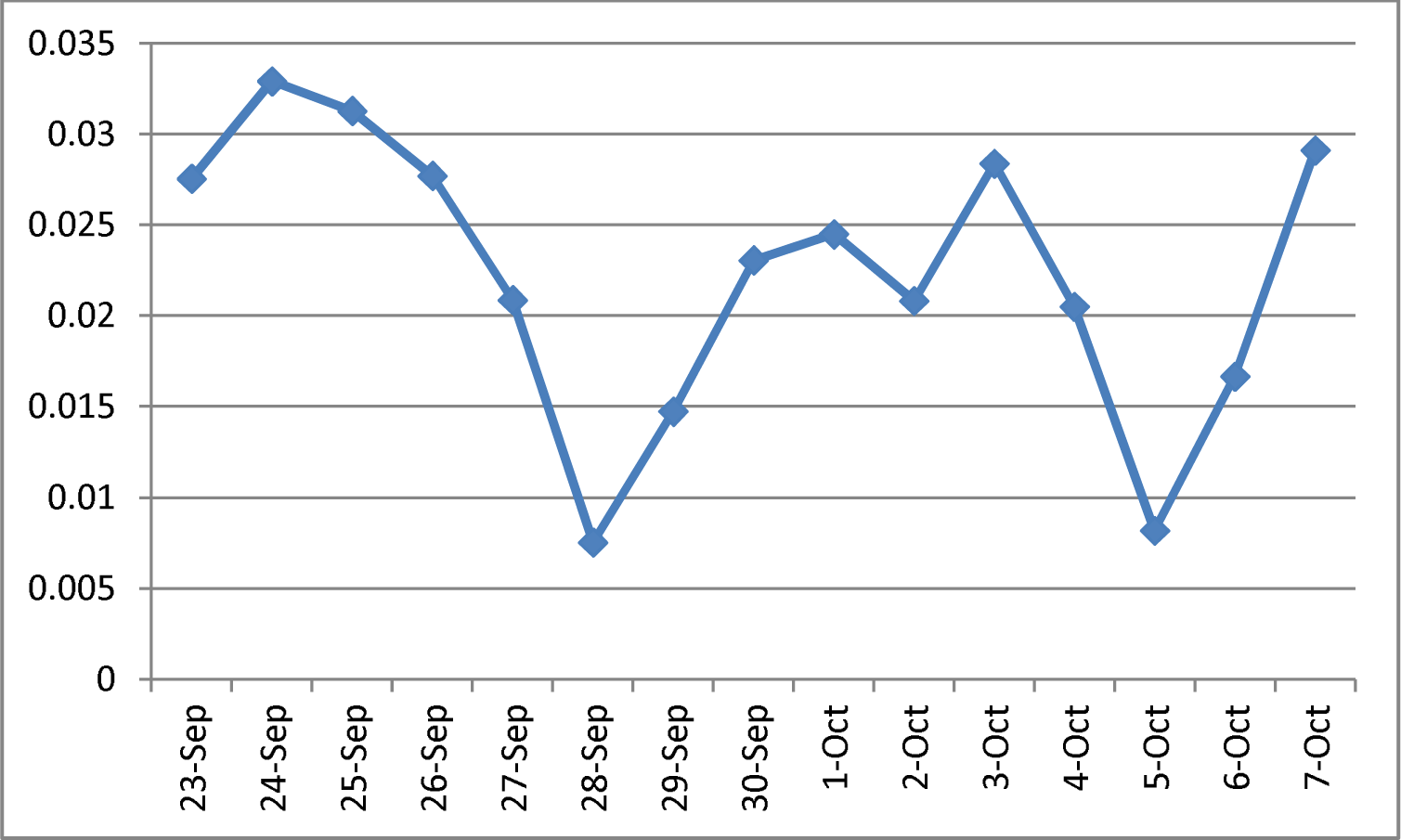
Values of Δ*z*(*t*), France, 23 September to 7 October.

It may be mentioned at this point that the troughs in the daily new cases [4] were seen to be minimum in Italy around June 27 to July 16, in Spain around May 20 to June 27, in France around June 8 to June 27, in the UK around 10 July, in Russia around August 23 and in Germany around June 8, and thereafter currently in October the disease has been spreading again, this time following very unusual patterns.

We shall now put forward a mathematical explanation of the unusual pattern that Δ*z*(*t*) has been following in Europe. When the epidemic had appeared, the cumulative total number *N*(*t*) of cases was following a nonlinear pattern. Soon the pattern became nearly exponential. In this nearly exponential phase *N*(*t*) was expressible approximately as

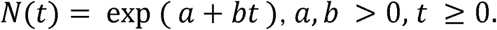

Defining

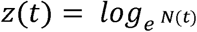

it was found in earlier studies [5, 6, 7, 8, 9, 10, 11] that the values of

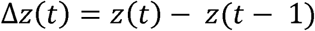

show a linearly reducing trend with respect to time until a retarding phase starts and in that phase the spread pattern becomes nearly logarithmic. In the standard epidemiological models also this kind of a growth pattern is assumed. However, the pattern in the second wave of the epidemic is different in the sense that Δ*z*(*t*) in these six European countries has been showing an increasing trend instead of a decreasing trend as was observed in earlier studies. There can be only one explanation of this phenomenon. In the first wave, *z*(*t*) was of first degree but in the second wave, whatever might be the reasons, *z*(*t*) is *not* of first degree. What we mean is that in this second wave the natural logarithm of *N*(*t*) is not a first degree polynomial in t, and that is why our calculated values of Δ*z*(*t*) are showing an increasing trend instead of a decreasing trend in Italy, Germany, Russia and the UK. In Spain and France, an increasing trend in the values of Δ*z*(*t*) is not clear yet, but the values are not showing near constancy also. Therefore it is obvious that in these two countries also *z*(*t*) is not of first degree in t. If in the second wave of the epidemic

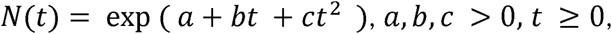

then it is obvious that Δ*z*(*t*) would show an increasing trend because *c* > 0, but this kind of an exponential growth was not seen in the first wave which is why it was observed that the values of Δ*z*(*t*) were showing a linearly decreasing trend earlier.

Therefore we can empirically infer that as long as Δ*z*(*t*) was an estimate of the coefficient b for some t in *N*(*t*) = exp (*a* + *bt*),*a,b* > 0, *t* ≥ 0, it was showing a decreasing trend in the nearly exponential phase of growth of the epidemic when the virus first appeared. Currently in the resurgence, *N*(*t*) is the exponent of a nonlinear function of time and therefore we have found the behavior of Δ*z*(*t*) unusual. In effect, the resurgence is following a much faster growth pattern different from what was seen to be followed when COVID-19 had appeared.

## Conclusions

The COVID-19 situation in Europe is showing an unusual trend. How long would the situation remain in this way cannot be said now. But one thing can be said with certainty that the situation has become explosive. In Russia in particular, the growth is not following the standard epidemiological growth pattern. In Italy, Germany and the UK too, the growth is very unusual. In Spain and France, the situation is uncertain as yet. We conclude that the growth pattern of the epidemic in the current resurgence in the six European countries concerned is of a type very different from the nearly exponential type followed in the accelerating phase in the first wave.

## Data Availability

The data were collected from Worldometers.info.

https://www.worldometers.info

